# The potential for digital patient symptom recording through symptom assessment applications to optimize patient flow and reduce waiting times in Urgent Care Centers: a simulation study

**DOI:** 10.1101/2020.06.19.20135590

**Authors:** Maryam Montazeri, Jan Multmeier, Claire Novorol, Shubhanan Upadhyay, Paul Wicks, Stephen Gilbert

## Abstract

**Background:** Overcrowding can negatively affect the performance of the health care facilities not only for patients in terms of delayed care delivery and increased health risk, but also for health care workers in terms of increased burden and stress. Sometimes overcrowding is a result duplicate activity such as history taking and recording of patients’ symptoms. In this case, using a digital symptom assessment application can prevent duplication of such activities and may decrease the crowding in health care facilities.

**Objective:** We sought to understand the effect of a digital symptom assessment app that facilitates the taking of patient clinical history to optimize patient flow. We hypothesized that waiting times and crowding in an urgent care center could be reduced through the introduction of a digital history taking tool, and that this would be more efficient than simply adding more staff.

**Methods:** A discrete event approach was used to simulate patient flow in an urgent care center during a hypothetical 4-hour time window. The baseline case simulated a small center with 2 triage nurses, 2 doctors, 1 treatment/examination nurse and 1 discharge administrator in service. In addition to the base case, the center is simulated in 32 scenarios either with different number of staff or different assumption on time saved by the app. Target outcomes included average queue length, waiting time, idle time and utilization of staff

**Results:** Discrete event simulation found that a few minutes saved by a digital history taking app during triage could significantly increase efficiency. An estimated time-saving per patient of 2.5 minutes decreased average patient wait for triage by 26.17%; a 5 minutes time-saving would lead to a 54.88% reduction. Alternatively, adding an additional triage nurse was less efficient, as the additional staff were only required at the busiest times. While reduction in waiting time for triage was similar (approximately 50%) for either approach, adding a triage nurse reduced the median nurse utilization from 97% to 41%, while adding the tool resulted in median nurse utilization of 88%.

**Conclusions:** Digital history taking could result in substantial reduction in patient waiting time for triage nurses, which is associated with reduced patient anxiety, staff anxiety and improved patient care. Patient history taking could be carried out in waiting room (via a check-in kiosk or portable tablet computer) or out at home. This simulation has the potential to impact service provision and approaches to digitalization at scale.

## Introduction

### Background

Overcrowding in healthcare facilities occurs when the number of patients seeking care exceeds the care facility’s capacity in a given time period. Long queues of patients can lead to delayed care delivery, increased health risk for urgent cases, higher rates of hospital-borne infections, increased stress, and avoidable staff burden [1,2]. Overcrowding has also been associated with increased occurrence of preventable medical errors and with negative effects on clinical trial outcomes [3–5]. Health care system performance can be measured in terms of patients’ waiting time and quality of the service, amongst other variables such as cost [6]. One method that can help analyse the performance of the whole system is patient flow modelling, which can aid decision making in planning capacity, resources, and appointment scheduling [7].

Methods to improve the flow of healthcare delivery include eliminating unnecessary and duplicate activities, performing activities in parallel, and identifying alternative process flows [7]. “History taking” and recording of patients’ symptoms by skilled labor is an activity which is often duplicated during triage and treatment in both urgent care centers (UCC) and emergency departments (EDs)[8]. First, a triage nurse asks for symptoms and patient history to classify patients into different levels of severities. Then, the treating physician later repeats this process in more detail history to inform the next appropriate steps for determining potential diagnoses and treatments[8].

### Digital history taking

Were history taking to be performed by a digital symptom assessment application by patients in the waiting room, this might enable professionals to save time and treat more patients [9]. One such tool, Ada uses a probabilistic reasoning engine to collect demographic information, medical history, and symptoms. A previous usability study found that patients using Ada’s tools in a primary care waiting room reported them helpful and easy to use [10]. A clinical vignette study showed that Ada’s reasoning engine has similar levels of coverage, accuracy, and safety as human general practitioners [11]. This is important because a symptom recording tool must be able to ask appropriate and targeted questions on the wide range of symptoms with which patients can present in primary care. However, it remains unclear what potential benefits might be experienced in a more urgent setting.

### Urgent Care Centers

The term UCC can refer to several types of service including walk-in centers, urgent care centers, minor injury units and urgent treatment centers, all with different levels of service [12]. As modelled in this study, a typical UCC led by a physician (general practitioner), open every day of the week, equipped to diagnose and treat common ailments such as: sprains and strains, suspected broken limbs, minor head injuries, cuts and grazes, bites and stings, minor scalds and burns, ear and throat infections, skin infections and rashes, eye problems, coughs and colds, feverish illness, abdominal pain, vomiting and diarrhea, and emergency contraception. For example, in the United Kingdom, this type of unit referred to by the NHS as an “urgent treatment center”[13]. While most prior research on triage, waiting and consultation time distributions has studied primary health clinics [14–16] or the ED [17–20], relatively little has been reported about UCCs. We found only one study [21] that compared waiting and consultation times in UCCs and physician offices.

### System Simulation for Workflow Efficiency

In order to understand the potential benefits of the Ada tool we used a system simulation approach, widely adopted for the exploration of the effects of new digital health technologies or process changes in health care system flow and efficiency. We sought to address the hypothesis that waiting times and crowding in an UCC could be reduced through the introduction of a digital history taking tool, and whilst this could also be achieved through the addition of staff, that system efficiency would be greater with the use of the tool.

## Methods

### Simulation Development

The scenario simulated in this study was based on current real-world use of intelligent digital symptom and history taking applications. We compared a scenario in which there was no digital symptom assessment to a scenario in which *every* patient entering the UCC waiting room had used the symptom assessment tool. Patient usage could be either: (i) at home (using a web-page or phone application), (ii) using check-in kiosks in a co-located ED waiting room, before fast track redirection to the associated UCC; or, (iii) using check-in kiosks at the UCC. In each case it was assumed that the assessment report’s questions, answers, demographics, and symptoms would be entered into the UCC’s electronic medical record (EMR) as “handover”.

### Parameter Development – Clinical Setting

We simulated a UCC in the first four hours of its opening. In the first step of the patient journey (see Figure 1), a triage nurse assesses the symptoms of the patient. In the next step, the patient visits the doctor and either visits the examination/treatment room (with the probability of λ) or is discharged (with the probability of 1-λ). If a patient visits the examination/treatment room, he or she is either redirected to the doctor for further investigations (with the probability of ω) or discharged (with the probability of 1-ω). Triage duration, consultation duration, number of staff in-service and arrival rate of the patients affect the patient flow in the UCC. The baseline scenario of staffing of the UCC was based on professional experience of one coauthor (S.U.) and another colleague (A.B.) who have each worked for over 5 years in a combination of NHS general practices, UCCs and EDs. We assumed that there were two triage nurses, two doctors, one nurse for examination/treatment and one administrator responsible for discharge (Table 1). We simulated the effects of each patient using a symptom assessment app on queue size, waiting time for triage nurses, idle time and utilization of triage nurses and doctors. Waiting times were not available from the literature for UCCs so we extracted such data from the ED setting [18][17].

**Table 1.** Number of staff and average duration of patient-staff interaction in the baseline setting of the UCC.

| Baseline setting | Triage nurse | Doctor | Examination / treatment nurse | Administration staff |
| --- | --- | --- | --- | --- |
| Average duration of interaction (min) | 15 | 20 | 15 | 5 |
| Number | 2 | 2 | 1 | 1 |

**Figure 1.**
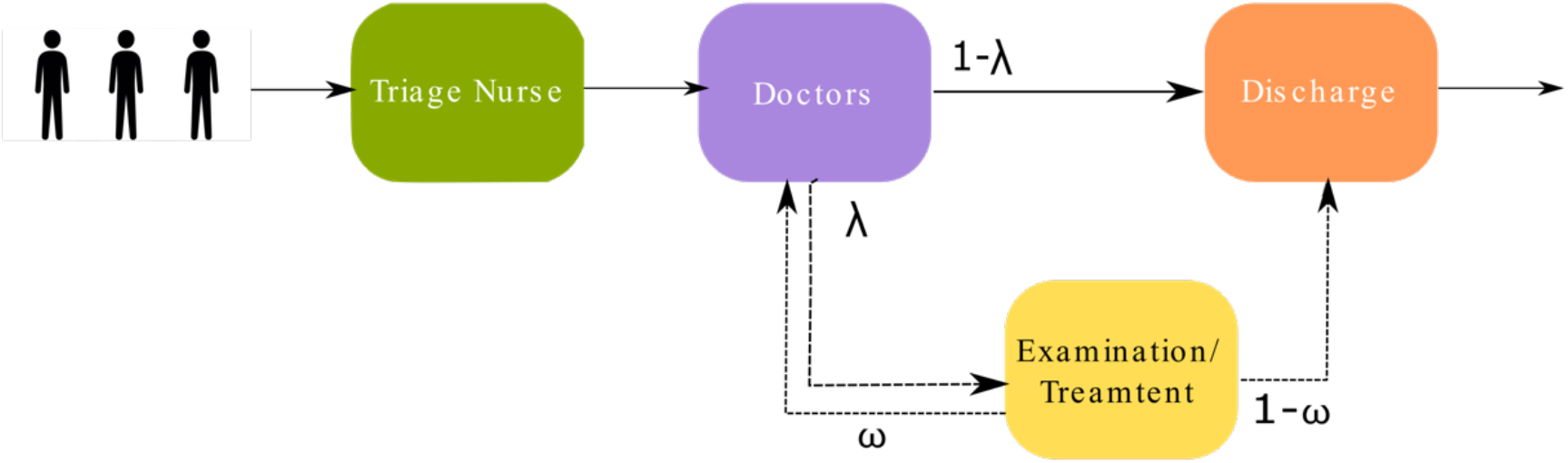
Illustration of the UCC, where patients arrive without any planned appointment. In the first step a triage nurse runs a symptom assessment, then patients are directed to the doctor. Depending on their situation they may be examined/treated by another nurse and then discharged or sent back to a doctor or discharged immediately by administrative staff.

### Parameter Development – Time Savings

Our model required a parameter for how much time could be saved through digital history taking. A 2017 pilot implementation of a symptom app assessment in a busy 10,000 patient UK primary care practice saved an estimated 1.9 minutes, as reported by doctors from over 300 primary care consultations (unpublished data). A 2019 pilot conducted structured interviews with 5 ED clinicians, who viewed the handover report produced by the app and estimated a time saving of between 4-6 minutes (unpublished data). Finally, in an observational study time savings in the ED were estimated in the range of 2.5-5 minutes by triage nurses and physicians [9].

### Setting Model Parameters - Overcrowding

Firstly, we simulated the flow with different arrival rates to cause overcrowding, defined as more than 5 patients waiting for staff. We simulated the patient trajectory starting with an arrival rate of 0.1 patients per minute. As the stability of the results depends on the number of simulations, we measured the queue sizes in different runs of simulation, i.e., 500, 1000, 5000 and 10000 in the baseline setting to find out how many simulation-runs lead to stable results. We found that after 5000 runs of simulations, the results did not change. For the arrival rate of 0.1 patients per minute, we observed an average queue size of around 1.27 and 2.43 patients for triage nurse and doctors respectively. As we increased the patient arrival rate from 0.1 to 0.2 patients per minute, we observed an increase in nurse queue size from 1.27 to 8.46 patients and a corresponding increase in doctor queue size from 2.43 to 5.30. Therefore, for further analysis, we used the arrival rate of 0.2 patients per minute, or one new patient every 5 minutes.

To explore our hypothesis that crowding can be reduced either through the addition of a digital tool, we simulated different *what-if-scenarios*. We also varied staff numbers from the baseline case scenario to explore staff utilization, as our hypothesis also recognized that crowding can likely be reduced by provision of more staff. We investigated the effect of changing the number of staff and using a symptom assessment app on queue status, waiting time for triage nurse, idle time and utilization of triage nurses and doctors. Here the waiting time for triage is defined as the interval between the time patient arrives at the triage nurse and the time patient leave the triage nurse excluding the action time which here meant as the time spent for triage. Idle time is defined as the duration when the number of occupied staff is less than their capacity, i.e. at least one member of the staff is not occupied. Utilization is calculated originally in the *simmer* package and is defined as the total time staff member are in use divided by the total time of simulation.

### Statistical Analysis

We used the package *Simmer* (version 4.4.0) [22] which is process-oriented and trajectory-based Discrete-Event Simulation (DES) package for R. Based on the observational study and clinician interviews, we therefore used a range of possible time savings (2.5 to 5 minutes) to parameterize the model (Table 2). In total, we simulated 33 different scenarios including the baseline setting, adding new staff and different combinations of time saved by the app for doctors and triage nurses to measure nurses’ and doctors’ queue sizes, their idle time and utilization and also waiting time for triage nurses. Each scenario was simulated 5000 times and the result metrics are reported as the overall mean and 95% confidence interval in 5000 runs of simulations, except for utilization which is originally calculated in R’s simmer package as the median, 25%, and 75% quartiles of 5000 runs of simulation. The baseline case scenario was an UCC staffed with two triage nurses, two doctors, one treatment nurse and one administrator responsible for the discharge. We assumed that patient arrivals, triage, consultation and discharge (all the events in patient flow through the UCC) follow a Poisson distribution. Therefore, in our simulations, the time interval distribution between all the events follow exponential distributions.

**Table 2.** The model was parameterized with a range of estimates of time savings for the triage and consultation processes, resulting in 30 different scenarios for the digital symptom and history taking tool.

|  |  |  |  |  |  |  |
| --- | --- | --- | --- | --- | --- | --- |
| **Time saved by app for triage, minutes** | 2.5 | 3 | 3.5 | 4 | 4.5 | 5 |
| **Time saved by app for consultation, minutes** | 1.5 | 2 | 2.5 | 3 | 3.5 | - |

## Results

Table 3 shows how extra staff and use of a digital tool could affect the overcrowding of patients in terms of queue size, idle time and utilization of staff and waiting time for triage nurse in comparison to the baseline case scenario.

**Table 3.** The effect of adding extra staff or using a digital symptom assessment app on the queue sizes, idle time, and utilization of staff members and patients’ waiting for the triage nurse.

| Scenario | Mean Queue size for triage nurses, # of patients (95%CI) | Mean Queue size for doctors, # of patients (95%CI) | Mean idle time of triage nurses, minutes (95%CI) | Mean idle time of doctors, minutes (95%CI) | Median utilization of triage nurses, % (25%-75% quantile) | Median utilization of doctors, % (25%-75% quantile) | Mean waiting time for triage nurses, minutes (95%CI) |
| --- | --- | --- | --- | --- | --- | --- | --- |
| Baseline | 8.47<br>(8.44-8.49) | 5.44<br>(5.42-5.46) | 13.99<br>(13.50-4.50) | 24.10<br>(23.40-24.80) | 96.9<br>(92.8-98.9) | 93.3<br>(86.9-97.1) | 34.05<br>(33.90-34.21) |
| Baseline+ triage nurse | 3.4<br>(3.37-3.39) | 9.53<br>(9.51-9.56) | 61.04<br>(59.86-62.22) | 13.43<br>(13.00-13.86) | 40.5<br>(24.9-62.9) | 96.1<br>(93.3-98.3) | 13.2<br>(13.13-13.28) |
| Baseline+ triage nurse+ doctor | 3.47<br>(3.46-3.48) | 5.57<br>(5.55-5.58) | 59.78<br>(58.63-60.94) | 40.34<br>(39.55-41.13) | 47.3<br>(66.2-31.3) | 90.2<br>(84.8-94.5) | 13.54<br>(13.46-13.62) |
| Baseline+ digital tool (assuming minimum minute time saving) | 6.29<br>(6.27-6.31) | 6.82<br>(6.80-6.84) | 22.74<br>(22.05-23.43) | 19.32<br>(18.72-19.92) | 94.4<br>(88.4-98.2) | 94.4<br>(89.5-97.6) | 25.44<br>(25.32-25.56) |
| Baseline+ digital tool assuming maximum time saving) | 3.84<br>(3.82- 3.85) | 8.2<br>(8.17- 8.22) | 22.74<br>(40.80- 42.6) | 17.04<br>(16.54- 17.53) | 88.0<br>(79.1-94.7) | 95.1<br>(90.5-97.9) | 15.5<br>(15.48-15.64) |

### Effect of additional staff

Addition of an extra nurse reduces the queue length for triage nurses around 60% but leads to about 75% increase of the queue length for the doctors. Providing one additional doctor can reduce the number of patients waiting for doctors to a similar situation as the baseline case (Figure 2A). As shown in Figure 3A, adding one extra triage nurse results in a 336% increase of triage nurses’ idle time and a 44% decrease of doctor’s idle time as more patients would be transferred to consultation in a shorter time. Adding one extra doctor led to a 67% increase of the mean idle time of doctors. (Table 3 and Figure 3A). In the baseline case, the median triage nurses’ utilization is 96.9%. Adding one extra triage nurse reduced this value to 40.5% (Figure 4A). The median utilization of doctors always maintains a utilization level of 90% or above. Figure 5A shows that adding a triage nurse leads to 61.23% reduction in the average waiting time for triage nurses.

**Figure 2.**
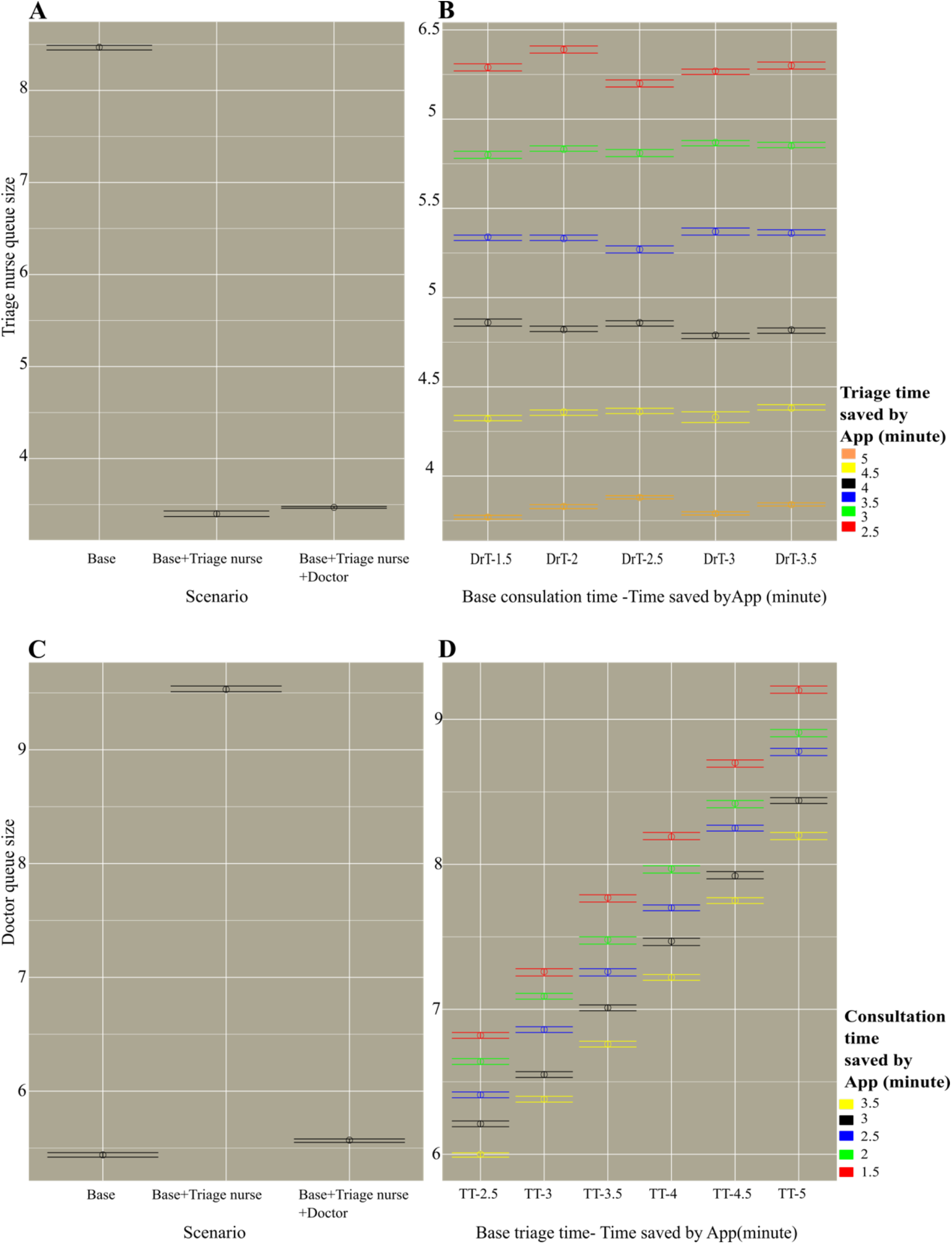
Queue size of triage nurses (A and B) and doctors (C and D). A and C represent the queues size of mentioned staff in scenarios where no App is used, which are base case setting and scenarios with extra staff. B and D on other hand, indicate queue sizes of triage nurses and doctors in 30 different scenarios of combining different time saving by app in triage and consultation process. In B, X axis labels show consultation time as a base doctor’s consultation time (DrT) subtracted by the time saved by App (1.5, 2, 2.5, 3, 3.5 minutes). In D, X axis labels show triage time as a base triage time (TT) subtracted by time saved by App (2.5,3,3.5,4,4.5,5 minutes). Each circle represents the mean value of 5000 runs of simulation in each of 32 combinations. Horizontal lines represent 95%confidence intervals of corresponding average.

**Figure 3.**
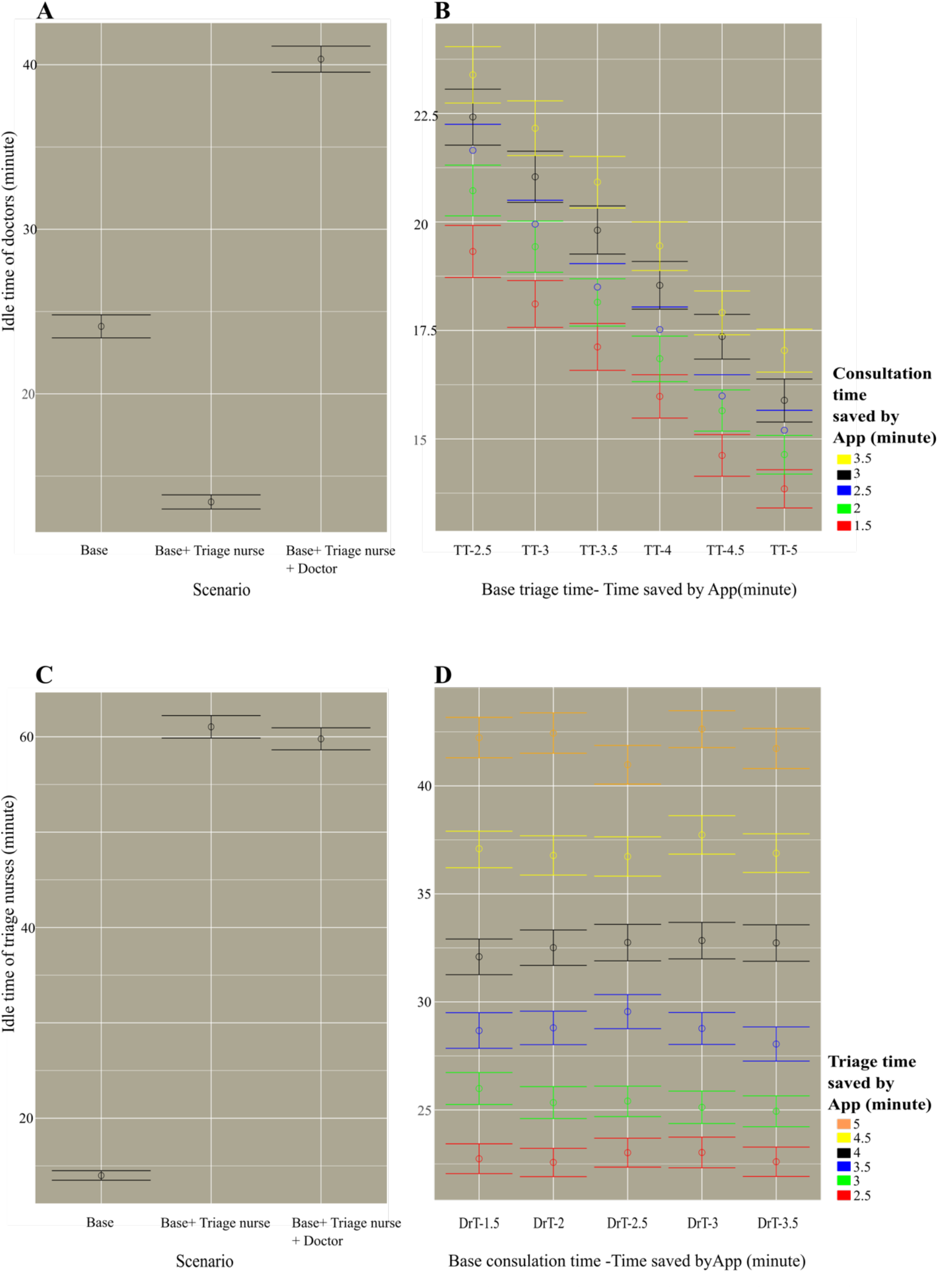
Idle time of doctors (A and B) and triage nurses (C and D). A and C represent the idle of mentioned staff in scenarios where no App is used, which are base case setting and scenarios with extra staff. B and D on other hand, indicate idle time of triage nurses and doctors in 30 different scenarios of combining different time saving by app in triage and consultation process. In B, X axis labels show triage time as a base triage time (TT) subtracted by time saved by App (2.5,3,3.5,4,4.5,5 minutes). In D, X axis labels show consultation time as a base doctor’s consultation time (DrT) subtracted by the time saved by App (1.5, 2, 2.5, 3, 3.5 minutes). Each circle represents the mean value of 5000 runs of simulation in each of 32 combinations. Horizontal lines represent 95% confidence intervals of corresponding average.

**Figure 4.**
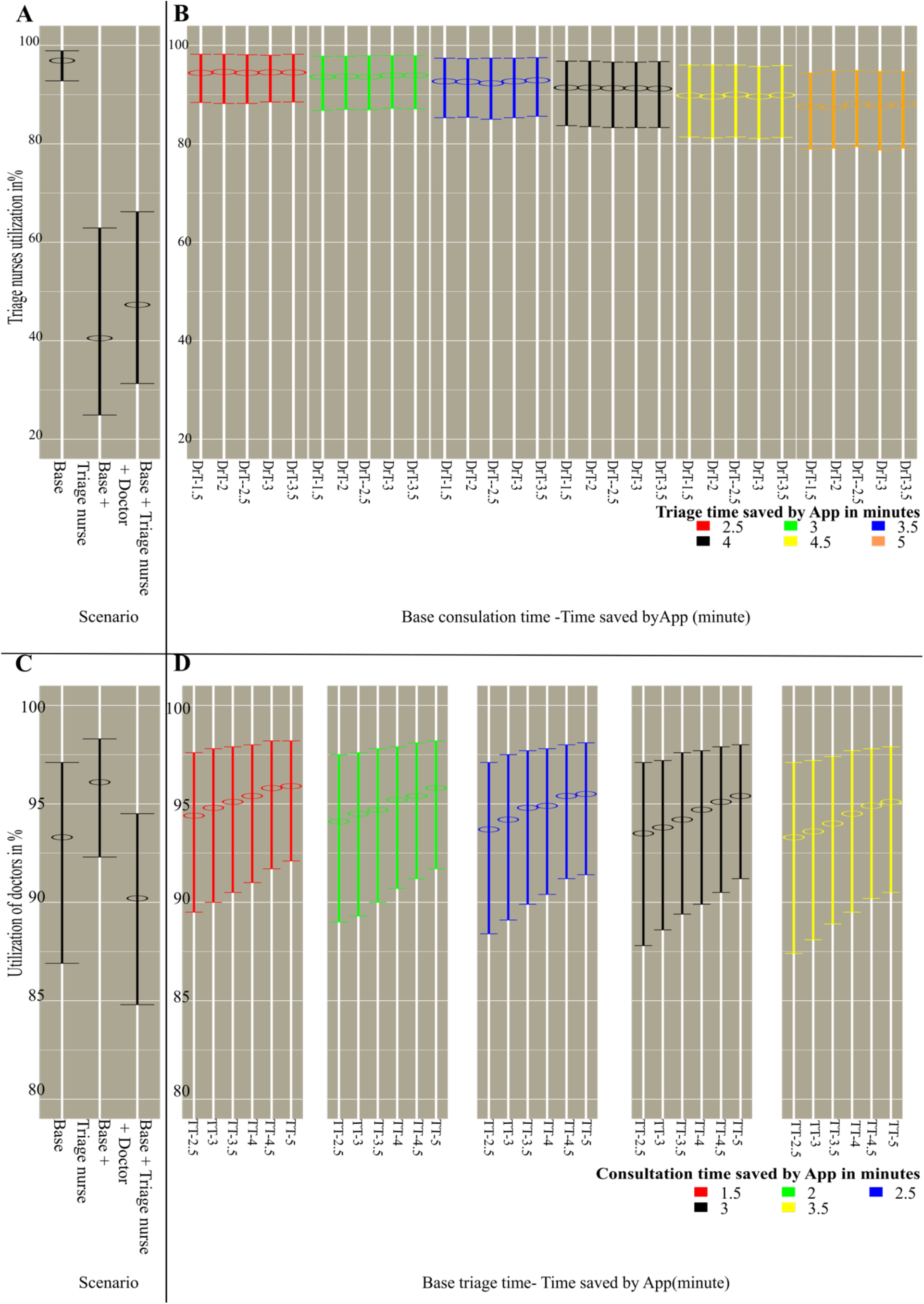
Utilization of triage nurses (A and B) and doctors (C and D) in percentage. A and C represent the idle of mentioned staff in scenarios where no App is used, which are base case setting and scenarios with extra staff. B and D on other hand, indicate idle time of triage nurses and doctors in 30 different scenarios of combining different time saving by app in triage and consultation process. In B, X axis labels show triage time as a base triage time (TT) subtracted by time saved by App (2.5,3,3.5,4,4.5,5 minutes). In D, X axis labels show consultation time as a base doctor’s consultation time (DrT) subtracted by the time saved by App (1.5, 2, 2.5, 3, 3.5 minutes). Each circle represents the median value of 5000 runs of simulation in each of 32 combinations. Horizontal lines represent 25% and 75% quantile corresponding average.

**Figure 5.**
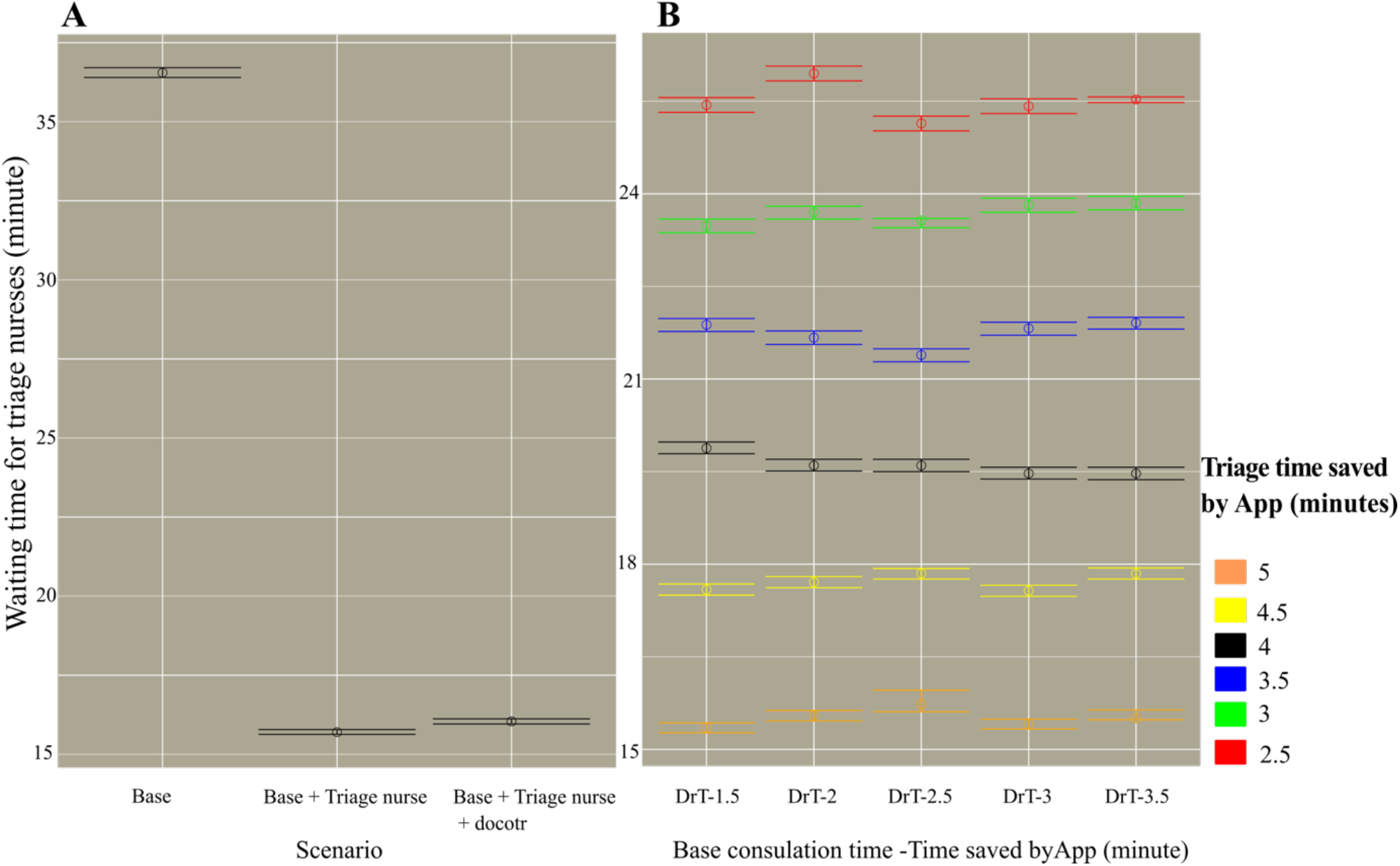
Waiting time for triage nurses in minutes. A represents the waiting time in scenarios where no App is used, which are base case setting and scenarios with extra staff. B on other hand, indicates waiting time in 30 different scenarios of combining different time saving by app in triage and consultation process. In B, X axis labels show consultation time as a base doctor’s consultation time (DrT) subtracted by the time saved by App (1.5, 2, 2.5, 3, 3.5 minutes). Each circle represents the mean value of 5000 runs of simulation in each of 32 combinations. Horizontal lines represent 95% confidence intervals of corresponding average.

### Queue sizes for triage nurses and doctors

Figure 2 indicates the results of applying a symptom assessment app on the queue sizes of triage nurses and doctors in comparison to baseline setting and addition of extra staff. Where time saved was assumed to be 5 minutes, the time-saving effect of digital symptom assessment on triage queue size was as great as adding one triage nurse. Even assuming a lower average time saving of 2.5 minutes per patient, this reduced the queue for triage nurses by 26.80%. However, when nurse-led triage takes less time per patient, then the rate of flow to doctors is increased, with a consequent increase in doctors’ queue size (Supplementary Table 1).

### Idle time of triage nurses and doctors

As shown in figure 3B, longer triage times and lower consultation times lead to longer idle times for the doctors. Assuming maximum app time saving, triage nurses’ idle time almost doubled whereas doctor idle time was reduced by 30%. By contrast, assuming minimum app time saving, average triage nurses’ idle time increased about 62% and average doctors’ idle time reduced by less than 20%. (Table 3, Supplementary Table 2)

### Utilization of doctors and triage nurses

Median triage nurses’ utilization dropped only moderately, about 9%, under maximum time savings. For minimum assumed time saving, this reduction was just 2.5% less than the baseline. Conversely the median utilization of doctors is less dependent on the amount of time saved and is always above 93%. (Figure 4, Table 3).

### Waiting time for triage nurses

Figure 5B shows that the more time saved by the app, the less time the patient waits for a triage nurse. For maximum time saving assumed, waiting time for a triage nurse was reduced by 54.88%. (Table 3, and Supplementary Table 4). Assuming minimal time saving, the waiting time for triage dropped by 25.28%.

## Discussion

### Principal findings

We simulated patient flow of an UCC in three conditions: a) baseline, b) with extra staff and c) with digital symptom taking. The shortest queue size and waiting time for triage nurses was achieved with the provision of one extra triage nurse (i.e., a total of three triage nurses) and one additional doctor (i.e., a total of two doctors). However, this approach may not be feasible due to limitations of available staff and high costs. Therefore, we hypothesized that the use of a digital symptom assessment app before the triage process could be another possible solution. Digital symptom assessment apps have the potential to improve the patient flow in health care facilities such as hospitals, primary clinics, EDs, and UCCs [23], where a long queue of patients not only puts a lot of pressure on the health care workers, but also on patients.

Our results suggest that for all measured variables, the amount of time saved by the app is an important determinant of the patient waiting time and system efficiency improvement. We found an amplification of time efficiency, through which relatively modest time savings per patient consultation accreted into substantial reduction in queuing time systemically. The minimum assumption of a 2.5-minute time saving from digital symptom taking reduced the patient waiting time for triage by 26.17%. This effect was even greater for the maximum assumption of saving 5 minutes per patient triage, which led to a 54.88% reduction in patient waiting time for triage nurse.

While overcrowding can be resolved by additional staff, simulation suggested that simply adding triage nurses may be inefficient as additional staff are only required at the busiest times. A digital symptom tool that could save 5 minutes per patient led to a reduction in waiting time equivalent to employing one extra triage nurse. Adding a triage nurse would have lowered staff utilization from 88% to 40%.

### Simulation for improving health system efficiency

Simulation is an accepted and powerful method for hypothesis generation for the effects of new healthcare interventions on overall system efficiency. Results of many simulation-based studies have already been implemented in real-world settings for better management of patient flow. One example evaluated scheduling, process flow and resource levels in an oncology center [24], where the implementation of the changes proposed by the simulations resulted in the improvement of the center’s system-wide performance. Another example applied the techniques explored here to a military outpatient primary care clinic. Simulation revealed a hybrid appointment/walk-in model for improving patient flow and optimized care provider utilization [25]. A final example applied a simulation model to identify factors contributing to flow blockage in an outpatient clinic of the Indiana University Medical Group. The strategic recommendations proposed from the simulation study led to significant improvements in real-life patient waiting time and physicians utilization [26].

There are two general methods to solve patient flow problems: i) queuing analytics and ii) simulation approaches. In the queuing theory approach, systems transition between a number of states, and are therefore most applicable to limited number of simplified models and for systems which will eventually reach a steady state[7,27]. By contrast, simulation approaches have fewer assumption and are therefore more flexible and versatile. Simulation methods such as system dynamics, agent-based simulation, and discrete event simulation have gained a lot of attention as a helpful method to tackle the complexities of analysis of patient flow in different areas. These applications include: (i) the detection of bottlenecks of the patient flow in healthcare facilities; (ii) optimizing flow management strategies such as scheduling and resource allocation rules; and, (iii) estimating treatment cost in terms of the lengths of stay of patients [7,28,29]. Here we used discrete event simulation, where patients are considered as independent entities [30] interacting with staff, such as nurses and doctors, through events like arrival, admission, and discharge.

### Comparisons to the wider literature

One of the principal reasons patients choose UCCs is that perceived waiting times are lower than in GP practices or in the ED [33]. However, we were unable to identify any time series studies that report waiting times or other clinical processes in UCCs, and there has been little systematic data gathering on UCC clinical efficiency [12]. There is more substantial health service delivery and clinical efficiency research on the ED setting [34], and although time series studies have been carried out, it is not reported with certainty how long the taking and recording of clinical history and symptoms takes, nor how much time can be saved through digital history taking tools. Although we found no studies investigating the benefit or performance of self-assessment with a digital assessment tool in the UCC, there are some studies reporting self-triage as potential for optimizing flow in subsections of Emergency or in primary care units. Amresh and Sinha[35] investigated the pediatric emergency department use of a kiosk bilingual self-triage system and concluded that kiosk based triage enabled parents to provide history faster and more accurately than routine nurse-initiated triage.

### Unanswered questions and future research

It is widely recognized that many promising digital innovations in healthcare are ultimately not adopted in practice, or are abandoned soon after limited local pilot utilization [37]. Often it is not the limitations of technology, or difficulties in implementation that ultimately determine the success of the pilots and wider adoption, but rather the dynamic interactions between many of these factors [38]. This study explored the potential effects of patient digital symptom and history taking on patient flow and queuing but does not explore the wider implications of the technology for the quality of care delivery, of patient experience, of patient safety or of the working experience of health care staff. These interlinked phenomena will be addressed in future studies.

ED overcrowding is mainly caused by patients who do not require urgent treatment [5] but whose medical history must be documented, accounting for some 41% of ED doctors’ time [8]. Overcrowding also leads to interruptions which impair history taking and documentation, particularly for inexperienced junior physicians who are overstretched [5].

Future research (including simulation studies, clinical investigations and technology role-outs) should seek to understand the potential of such tools in reducing documentation burden, facilitating fast tracking, increasing patient safety, improving documentation accuracy, and ultimately reducing overcrowding.

### Strengths and limitations of this study

We used discrete-event simulation to simulate a queue of events. The choice of modeling technique, model structure, and chosen parameter values limit the generalizability of results as the nature of UCCs varies substantially [12]. In our model we only considered a UCC without any planned appointments. We also assumed a first-in-first-out flow, irrespective of the urgency of treatment of individual patients. Patients and staff were all treated as passive and we did not consider any ongoing learning that can influence patients and health worker interactions. We also assumed that there were enough digital devices available such that digital symptom assessment would not itself lead to another queue.

We modelled with the assumption that time spent taking history taking time leads to a time saving for both the triage nurse and for the treating physician. One example from the literature highlights the level of duplication in a typical ED setting [8], where a history was taken by: (i) the triage nurse; (ii) clerking (student) physician; (iii) 2nd-clerking on transfer to the acute medical unit; (iv) at history review in the general ward round; (v) at history retaking on admission to a specialty ward. In the ED setting the retaking of clinical history provides no clinical benefit, with the history often recorded near verbatim to the previous history, as part of a recognized ‘futile clinical cycle’ [8]. However, we acknowledge that sometimes the histories by the triage nurse and the treating doctor have different purposes. We assumed here that the information queried, and the time spent in both cases overlapped to a large degree. Finally, we assumed that nurses and doctors could assess the recorded symptoms within their standard workflow.

## Conclusions

This simulation showed that even a small reduction in the time taken to assess symptoms can lead to a substantial reduction in the time patients wait for triage nurses, which could in turn lead to reduced patient anxiety, lower staff anxiety and improved patient care. Compared to baseline, the use of a digital symptom taking tool shortened the average patient waiting time to the same extent as adding an additional triage nurse to the UCC, with the additional advantages of higher staff efficiency. Such approaches have the potential to streamline service provision and accelerate approaches to digitalization in urgent care settings.

## Data Availability

All the data in this study, are shown in tables and figures. The simulation data will be delivered if asked.

## Acknowledgements

We are grateful to Adel Baluch (Medical Expert, Ada Health GmbH) for advice on the normal structure, layout and working practices of UCCs.

## Contribution statement

MM, JM, CN, SU & SG contributed to the planning (study conception, protocol development). MM carried out the simulation setup and simulations. MM, JM & SG contributed to the data analysis & interpretation. MM, JM, CN, SU, PW & SG contributed to the reporting (report writing). All the authors contributed to commenting on drafts of the report. SG is the guarantor for this work. The corresponding author attests that all listed authors meet authorship criteria and that no others meeting the criteria have been omitted.

## Abbreviations

ED: Emergency department
EMR: Electronic medical record
UCC: Urgent Care Centre

